# Magnitude and timing of the antiviral response determine SARS-CoV-2 replication early in infection

**DOI:** 10.1101/2021.01.22.21249812

**Authors:** Nagarjuna R. Cheemarla, Timothy A. Watkins, Valia T. Mihaylova, Bao Wang, Dejian Zhao, Guilin Wang, Marie L. Landry, Ellen F. Foxman

## Abstract

The interferon response is a potent antiviral defense mechanism, but its effectiveness depends on its timing relative to viral replication. Here, we report viral replication and host response kinetics in patients at the start of SARS-CoV-2 infection and explore the impact of these kinetics experimentally. In both longitudinal patient nasopharyngeal samples and airway epithelial organoids, we found that SARS-CoV-2 initially replicated exponentially with a doubling time of ∼6hr, and induced interferon stimulated genes (ISGs) with delayed timing relative to viral replication. Prior exposure to rhinovirus increased ISG levels and blocked SARS-CoV-2 replication. Conversely, inhibiting ISG induction abrogated interference by rhinovirus and enhanced SARS-CoV-2 replication rate. These results demonstrate the importance of initial interferon-mediated defenses in determining the extent to which SARS-CoV-2 can replicate at the start of infection and indicate that biological variables that alter the airway interferon response, including heterologous induction of innate immunity by other viruses, could profoundly impact SARS-CoV-2 susceptibility and transmission.

## Introduction

The novel coronavirus SARS-CoV-2 emerged in late 2019 and has led to a global pandemic, causing over 98M infections and 2.1M deaths at the time of this writing (Dong, 2020). This huge impact has motivated efforts to understand the host immune response to this virus, both to better predict patient outcomes and to design interventions. For an emerging viral infection such as SARS-CoV-2, innate immune responses can be particularly important in host protection, as these responses do not require prior exposure to effectively defend against a pathogen. Studies based on patient samples indicate that dysregulation of innate immune responses late in infection drives immunopathology in severe COVID-19 (Galani et al., 2021; Lee and Shin, 2020; Lucas et al., 2020), but there are relatively few reports describing host responses to SARS-CoV-2 in patients at the start of infection, when innate immune mechanisms are most likely to contribute to host defense.

SARS-CoV-2 enters the body and first replicates in the upper respiratory tract, achieving the highest viral load in the first few days following infection (Wolfel et al., 2020; Zou et al., 2020). High viral load in the nasopharynx correlates strongly with transmissibility in contact tracing studies, and significant viral replication following exposure is likely a prerequisite, although certainly not the only factor, for COVID-19 disease progression (Cevik, 2020; He et al., 2020). A likely candidate for controlling the infection at the earliest stages is the mucosal interferon response. This defense mechanism is initiated when pattern recognition receptors within epithelial cells and immune cells sense general features shared by many viruses, such as common structural features of viral RNA. This recognition event triggers expression of type I and type III interferons (IFNs) and interferon-stimulated genes (ISGs). Secreted interferons, in turn, bind to cell surface receptors on nearby cells, amplifying ISG expression and creating an antiviral state in the mucosal barrier. Many ISGs encode effectors which directly block viral replication within virus target cells, whereas others encode cytokines which recruit and activate cells of the immune system (Odendall and Kagan, 2015; Schneider et al., 2014).

Recent evidence supports a protective role for the interferon response in COVID-19, although there is also evidence that the virus antagonizes this response. Recombinant interferon blocks SARS-CoV-2 replication in vitro, and genetic deficiencies in the Type I interferon response as well as anti-interferon autoantibodies have been linked to greater COVID-19 disease severity (Bastard et al., 2020; Lokugamage et al., 2020; Pairo-Castineira et al., 2020; Vanderheiden et al., 2020; Zhang et al., 2020). Furthermore, early data from trials of recombinant Type I or Type III interferon for COVID-19 indicate a therapeutic benefit, particularly when patients are treated early in disease(Feld, 2020; Monk, 2020; Wang, 2020). However, during initial infection of the upper respiratory tract, the kinetics of ISG induction by SARS-CoV-2 are not clear. ISG expression in SARS-CoV-2 infected epithelia can be observed in vitro and in patients, but there is also strong evidence that SARS-CoV-2 antagonizes the interferon response, which likely affects the magnitude and timing of this response (Banerjee et al., 2020; Blanco-Melo et al., 2020; Konno et al., 2020; Martin-Sancho et al., 2020; Ravindra et al., 2020; Xia et al., 2020; Zhou et al., 2020). Since a major beneficial function of ISGs is preventing viral replication, the kinetics of the interferon response early in infection are likely to determine its protective impact, and host and environmental factors which modulate the timing of this response may be key determinants of whether the virus can amplify to a high viral load following infection.

One factor that can potentially modulate antiviral defenses in the airway epithelium is induction of the ISGs by other viruses, and such effects may be particularly important in limiting viruses which successfully block autologous interferon induction. Rhinovirus, the most frequent cause of the common cold, is frequently detected in the human upper respiratory tract in the presence and absence of symptoms, and both symptomatic and asymptomatic infections can induce ISG expression in the upper respiratory tract mucosa(Landry and Foxman, 2018; Wolsk et al., 2016; Yu et al., 2019). Therefore, rhinovirus is an example of a common environmental factor which could potentially alter the kinetics of ISG expression at the initial target site of SARS-CoV-2 infection, the airway epithelium.

Here, we studied the initial host response to SARS-CoV-2 infection and its relationship to viral replication, including modulation by rhinovirus infection. Due to SARS-CoV-2 screening and testing practices at our hospital at the start of the pandemic in March 2020, we were able to obtain nasopharyngeal swab samples from patients at different time points post-infection, including serial samples collected close to the start of infection in asymptomatic subjects. Using transcriptomic and biomarker-based analysis of these samples, we observed robust ISG induction in the airway mucosa in response to SARS-CoV-2, but a delay between viral replication and ISG induction. Using an organoid infection model, we modulated the kinetics of ISG induction. We found that enhancing ISG expression by prior exposure to a rhinovirus profoundly inhibited SARS-CoV-2 replication. Blocking ISG induction completely rescued the effects of interference by rhinovirus, and increased replication rate of SARS-CoV-2 in a low MOI infection. These results show the importance of interferon-mediated defenses in restricting SARS-CoV-2 replication at the start of infection and provide an example of how ISG induction by a different virus could impact susceptibility to SARS-CoV-2 infection.

## Results

### Nasopharyngeal host response to SARS-CoV-2 infection identified by RNA-Seq

The host response to SARS-CoV-2 in the upper respiratory tract and its relationship to viral replication are not well-defined. To characterize host responses during SARS-CoV-2 infection in vivo, we performed RNA-Seq on nasopharyngeal (NP) swab RNA from SARS-CoV-2-positive patients (n=30) and SARS-CoV-2-negative healthcare worker controls (n=8). Patients included outpatients and patients admitted to the hospital of both sexes, who ranged in age from 20s to 90s, with the majority above 60 years of age (Fig S1A-D). Samples varied in viral load over >5 orders of magnitude as assessed by RT-qPCR for the SARS-CoV-2 N1 gene or by read mapping of NP RNA to the SARS-CoV-2 genome, with a strong correlation seen between PCR Ct value for the SARS-CoV-2 N1 gene and viral RNA reads by RNA-Seq (r^2^=0.8380, p<0.0001; Fig S1E).

Of reads mapping to the human genome, 1770 RNAs differed significantly between SARS-CoV-2+ patients and control subjects (Fig 1A). These included 1567 protein-coding genes, of which 1245 (79.4%) were enriched and 322 reduced in patients relative to controls. The most significantly enriched genes in the nasopharynx of SARS-CoV-2 patients were known interferon stimulated genes (ISGs), including three NP ISG transcripts previously shown by our group to accurately identify patients with viral respiratory infection, OASL, IFIT2, and CXCL10 (Fig 1A)(Landry and Foxman, 2018). Analysis of ingenuity pathways and transcription factor binding sites associated with enriched transcripts demonstrated activation of multiple pathways related to ISG induction in SARS-CoV-2 patients compared to controls, as well as other pathways linked to innate immunity, leukocyte recruitment, and initiation of mucosal inflammatory responses (Fig 1B-D).

**Fig 1.**
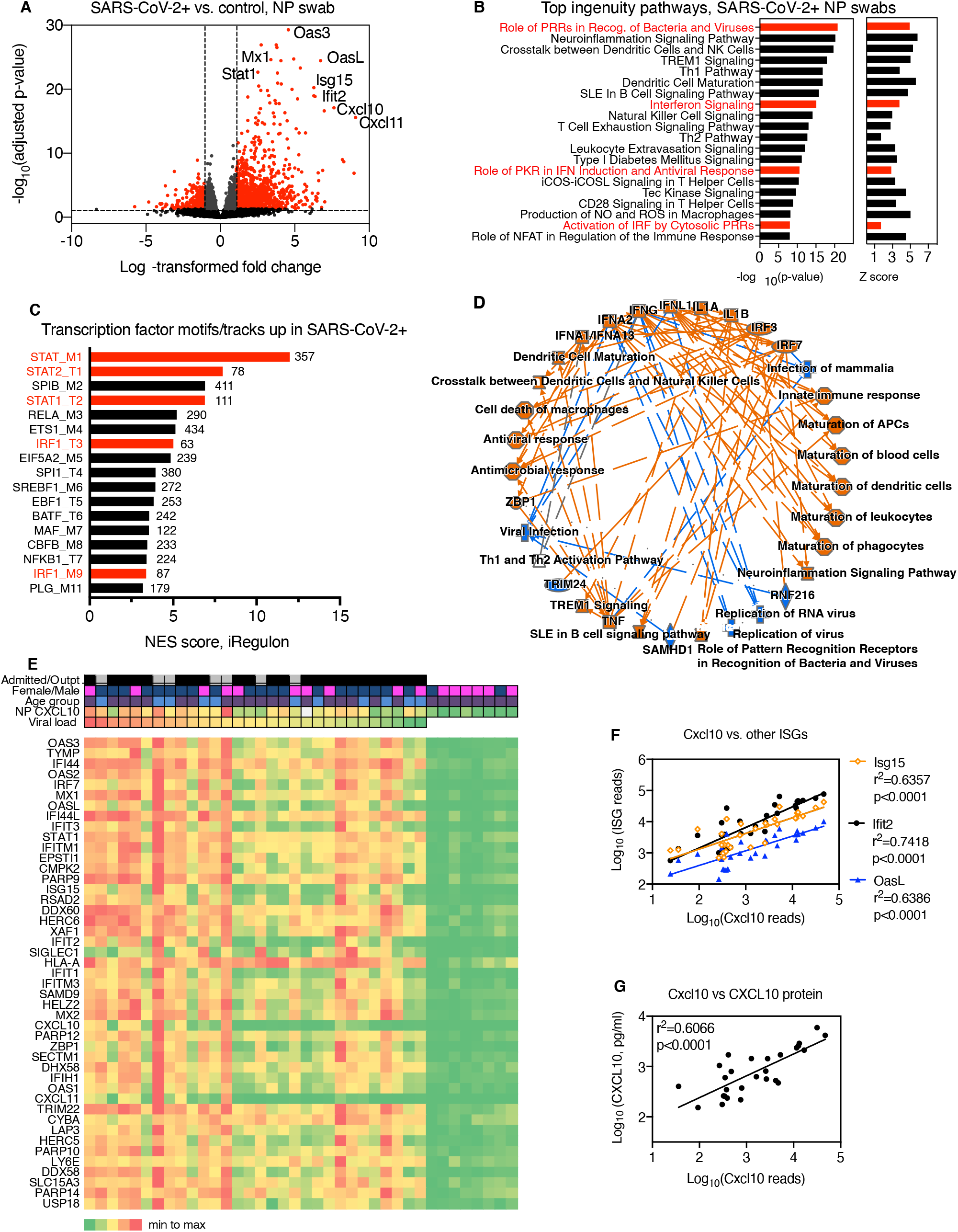
Transcriptome analysis of RNA isolated from SARS-CoV-2+ nasopharyngeal swabs (related to Fig S1). (A) Volcano plot showing significantly differentially expressed protein-coding genes based on RNASeq of NP swab RNA from SARS-CoV-2 patients (n=30) compared to control SARS-CoV-2 negative subjects (n=8). Transcripts with fold change>2, adjusted p-value<0.05 are highlighted in red. (B) Top 20 ingenuity pathways enriched in SARS-CoV-2+ compared to controls, based on 1770 differentially expressed RNAs. P-value and Z-score for each pathway is indicated on the x-axis. Pathways related to interferon and interferon regulatory factor (IRF) signaling are highlighted in red. (C) Transcription factor binding sites associated with NP transcripts enriched in SARS-CoV-2+ patients compared to controls. Bars show strength of association of motifs/tracks with enriched transcripts, indicated by NES score. Y-axis label indicates top transcription factor associated with each cluster of motifs (M) or tracks (T) and the cluster code. Number of enriched transcripts associated with each track/motif is indicated to the right of each bar. Transcription factors associated with the interferon response are highlighted in red. (D) Graphical summary of pathways and regulators enriched based on ingenuity pathway analysis of differentially expressed genes enriched in NP RNA of SARS-CoV-2+ patients compared to controls. (E) Heatmap showing relative expression level of top 45 most significant differentially expressed genes in patients (left) or SARS-CoV-2-negative controls (right). Clinical characteristics of each patient are indicated by color: viral load (red=highest viral load/lowest Ct value, green=lowest viral load/highest Ct value); NP CXCL10 protein level (red=highest, green=lowest, white=data not available). Heatmap colors represent values from highest (red) to lowest (green) for viral load (based on Ct value), CXCL10 concentration (pg/ml), or gene expression level, scaled from minimum to maximum (green=0; yellow=0.5, red=1) Patient characteristics indicated at the top of the graph include Admission status (grey=outpatient, black= admitted); Gender (blue=male, pink=female); Age (blue<55yrs, purple>60 yrs.) White = data not available. (F) Correlation between reads mapping to CXCL10 and reads mapping to other ISGs (Ifit2, OasL, Isg15). (G) Correlation between reads mapping to Cxcl10 and CXCL10 protein measured by ELISA in NP swab-associated viral transport medium.

Examination of gene expression across patient samples revealed several patterns (Fig 1E-G). First, the 45 most significantly enriched genes were all interferon stimulated genes, according to the Interferome database (Rusinova et al., 2013). ISGs appeared to be co-regulated within individual patients, i.e. patients with high expression of one ISG tended to have high expression of other ISGs (Fig 1E). This was also demonstrated by analysis of the correlation between reads for different ISGs across samples (Fig 1F). Second, ISG expression appeared to be loosely correlated with viral load, with those patients with the highest viral load (Fig 1E, left) tending to have higher ISG expression than those with the lowest viral loads (Fig 1E, right). However, while all SARS-CoV-2+ samples showed enrichment of ISG expression compared to controls, direct comparison of DEGs in patient groups with distinct clinical characteristics (sex, age, or outpatient/admitted status) showed no significant differences in ISG expression.

Next, we measured the level of CXCL10 protein in the NP-swab associated viral transport medium using ELISA. We previously showed that NP CXCL10 is detected in the viral transport medium during other acute viral respiratory infections and correlates with expression of ISGs at the mRNA level (Landry and Foxman, 2018). Consistently, we observed a significant positive correlation between NP CXCL10 protein level with the NP mRNA level of Cxcl10 in SARS-CoV-2+ patient NP samples (Fig 1E, G). Together, these results indicated that across subjects with diverse clinical presentations, SARS-CoV-2 induced a robust interferon response in the nasopharynx and that the NP CXCL10 protein level correlated with ISG expression at the RNA level.

### Relationship between SARS-CoV-2 viral load and NP host response in vivo

Next, we sought to examine the host response during SARS-CoV-2 infection in a larger set of samples from patients evaluated in our healthcare system in March and April of 2020 (n=140) (Fig 2 and Fig S2A, B), to gain further insight into the relationship between viral replication, disease status, and host response to infection. Based on our previous studies and our finding that NP CXCL10 protein level correlated with ISG expression in SARS-CoV-2 positive samples (Fig 1), we used NP CXCL10 as an indicator of the nasopharyngeal antiviral response. First, we examined NP CXCL10 level in SARS-CoV-2+ individuals who were tested as outpatients and not admitted to the hospital, compared to those who were admitted. Notably, we observed significantly higher NP CXCL10 levels in outpatients compared to admitted patients (Fig 2A, p=0.0019). To understand the reason for this, we first examined patient age, since as a group, admitted patients were significantly older than outpatients (16 years older on average, Fig S2C). However, there was no correlation between age and CXCL10 level (Fig S2D). Next, we examined viral load. We were initially surprised to find that admitted patients had significantly lower viral loads than outpatients (Fig 2B). This suggested that the main factor driving CXCL10 level was viral load. Supporting this idea, correlation analysis showed a significant positive correlation between NP CXCL10 level and viral load by RT-qPCR (for all patients, r^2^=0.2030, p<0.0001, Fig 2C). This correlation was also seen in separate analyses of outpatients and admitted patients, with but no significant difference in the slope of the CXCL10 vs viral load correlation between these groups, although there was a trend towards a higher slope in outpatients (Fig 2C). We also observed no significant relationship between sex and NP CXCL10 or sex and viral load in this sample set (Fig S2 E,F).

**Fig 2 (related to Fig S2).**
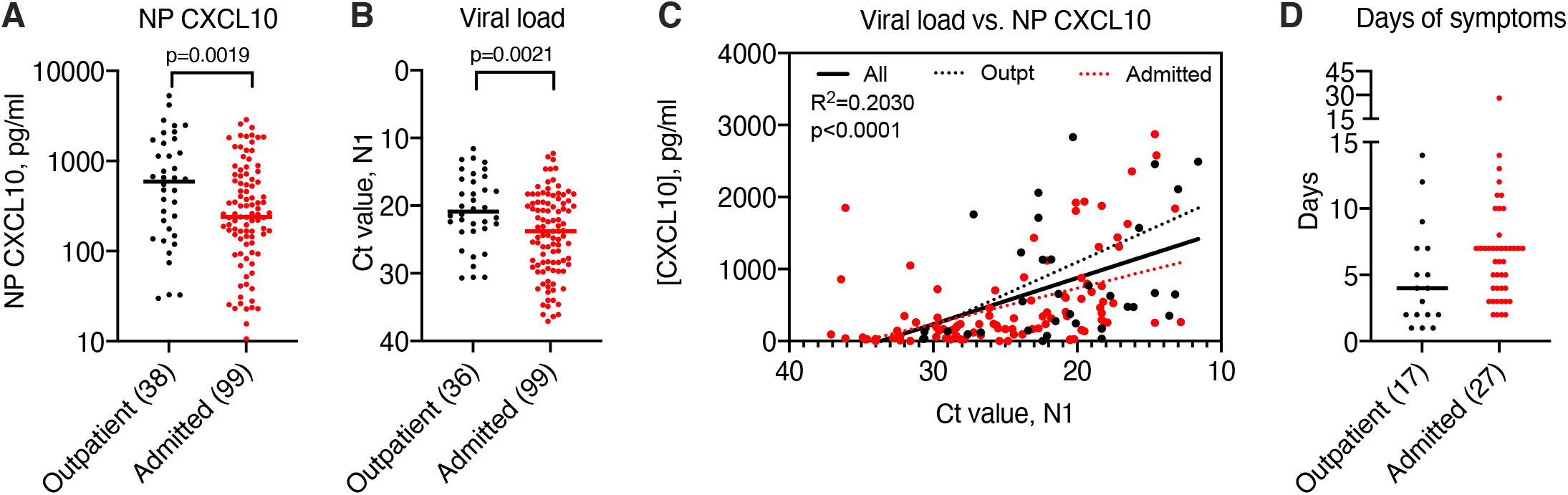
Relationship between nasopharyngeal (NP) CXCL10 and viral load in 140 SARS-CoV-2+ patients. (A) NP CXCL10 level in patients testing positive for SARS-CoV-2 by RT-qPCR at Yale-New Haven Hospital in March 2020. Black symbols indicate patients tested as outpatients or in the emergency department and not admitted to the hospital, red symbols indicate patients admitted to the hospital. (B) Ct value for SARS-CoV-2 N1 gene in outpatients and admitted patients. Number in parenthesis (A, B) indicates number of outpatient (total n=38) and admitted (total n=102) samples for which data was available. (C) Regression analysis showing relationship between viral load and NP CXCL10 protein for all samples (black solid line, r^2^ and p-value indicated) or for only outpatients (dashed black line) or admitted patients (dotted red line). (D) Days of symptoms reported prior to testing for samples with this information available (number indicated n parentheses).

Prior work on SARS-CoV-2 has shown that the nasopharyngeal viral load is highest in the first few days of infection, and that the more severe symptoms of COVID-19 requiring hospitalization occur in the second or third week of infection(Cevik, 2020). Therefore, we hypothesized that admitted patients may have shown lower viral loads at the time of testing than outpatients because they presented later in infection, after peak viral replication in the nasopharynx. Consistently, outpatients tended to report fewer days of symptoms prior to testing compared to admitted patients, although this information was only available for a subset of patients (about one-third, n=44; Fig 2D).

### Viral load and nasopharyngeal CXCL10 patterns in vivo over time

To further evaluate the relationship between viral replication and the innate antiviral response in the nasopharynx, we examined viral load and NP CXCL10 data in longitudinal samples. First, we examined viral load from 29 inpatients from March 12 and April 30, 2020 for whom we had at ≥8 sequential tests results for SARS-CoV-2 with at least the first sample tested using the CDC assay (our clinical laboratory also had other testing platforms) (Fig 3, Table S1). At this time, most serial testing was aimed at patient clearance for discharge. Consistently, the majority of patients (15/29) showed low viral loads (Ct N1>21) which remained low throughout the time course. Another common pattern was high viral load in the first sample (Ct N1<20) followed by a decline in viral load over time, similar to patterns reported in the literature for patients who presented close to the start of symptomatic illness (7/29 patients, Fig 3A). These patients showed high CXCL10 level in the sample with peak viral load and a decline in NP CXCL10 after the viral load had decreased (Fig 3B). One patient had a consistently high NP viral loads for 20 days and did not survive (not shown). Finally, a third pattern was seen in a several patients (6/29), in which the first sample had a low viral load which subsequently increased to a high peak level (Ct N1<20), then decreased over time (Fig 3). This pattern is consistent with patient presentation close to the start of infection. Two of these patients had no symptoms of SARS-CoV-2 and the virus was detected incidentally on screening during hospitalization for other reasons (Fig 3C,D, Table S1). One of these patients had an inconclusive test and a positive test on the same day, 12 hours later (first test, N1 not detected, N2 Ct 38.4, second test N1 Ct 34.6, N2 Ct 35.4), suggesting that this might have been the first day of infection for this patient (L2, Fig 3C). The other four patients presented with acute symptoms including fever, and in some cases cough and/or shortness of breath. NP CXCL10 in these patients was undetectable or low in the first positive sample with low viral load, then rose with viral RNA, and then subsequently declined as viral load declined. Together, the longitudinal data show a correlation with NP CXCL10 and viral load in individual patients over time, similar to what we observed across 140 patients tested at a single time point (Fig. 2). Notably, for patient L2, the only patient for whom three samples were available prior to peak viral load, there appeared to be a delay between CXCL10 production relative to viral replication during the first few days of infection, suggesting that initially viral replication outpaced the host innate immune response in the nasopharynx.

**Fig 3 (related to Table S1).**
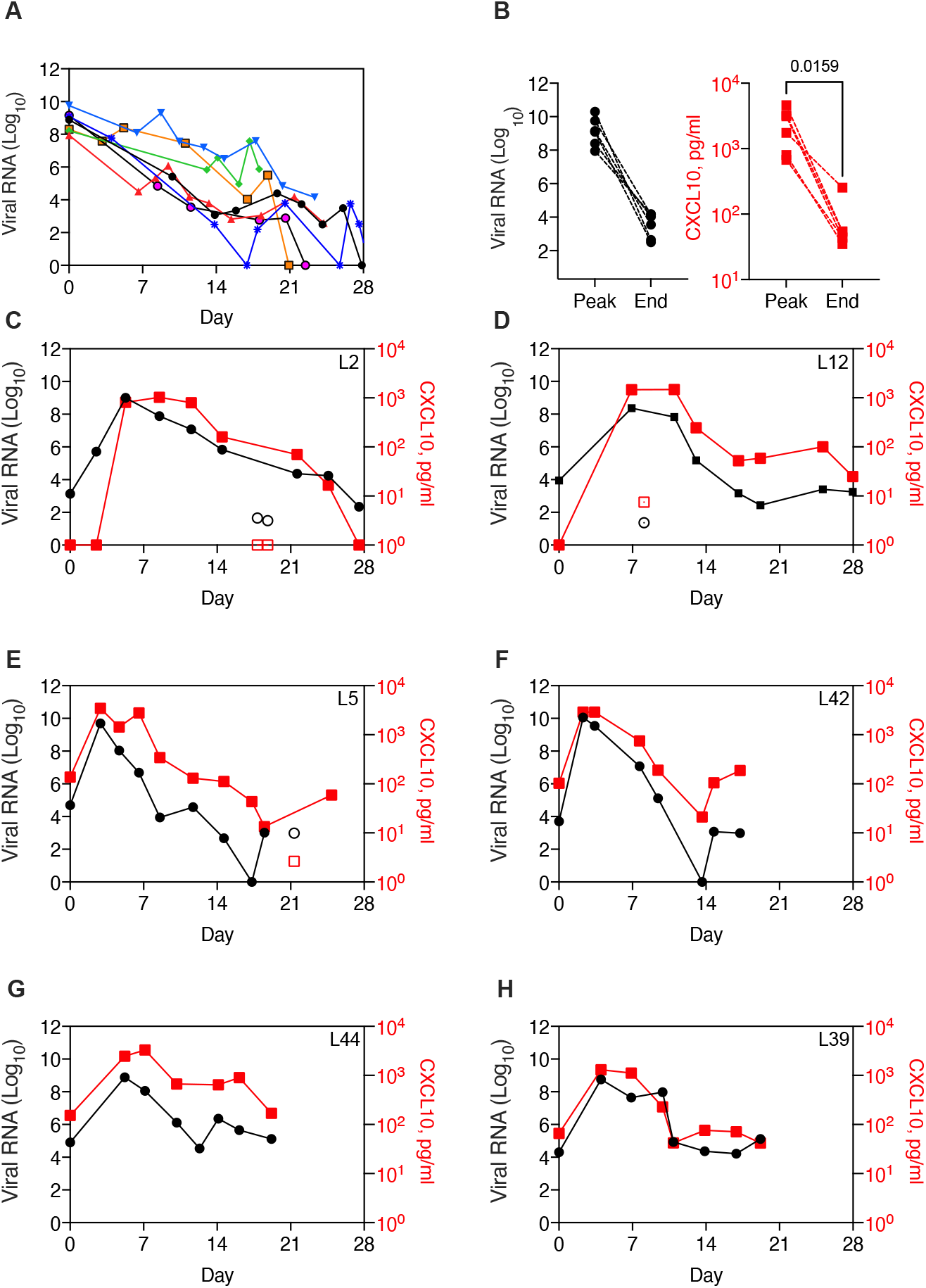
Nasopharyngeal viral load and CXCL10 in patients diagnosed prior to peak viral load. (A) Viral load over time in seven longitudinal samples from patients with high viral load in first sample (Ct N1>20). (B) Paired viral RNA and NP CXCL10 measurements at the peak viral load and at the end viral load, defined as the first sample with Ct N1>30, for 6 patients shown in (G) (data not available for one sample). CXCL10 level was significantly different in peak and end samples by paired t-test. (C) – (H) Viral load and NP CXCL10 level in longitudinal samples from SARS-CoV-2+ patients who presented with a low viral load (Ct N1>28) that increased to a high viral load (Ct N1<20). Viral load is expressed as fold change from the limit of detection for the SARS-CoV-2 N1 gene (black circles) and CXCL10 is expressed as pg/ml in the NP-swab associated viral transport medium (red squares). Samples with low levels of RnaseP, an indicator of sample quality, are shown with open symbols. Patient characteristics are described in Table S5.

### SARS-CoV-2 replication kinetics and ISG response early in infection

To further evaluate the kinetics of SARS-CoV-2 replication and host response, we performed a time course of SARS-CoV-2 infection using primary human airway epithelial cells grown at air-liquid interface, which differentiate into organoids with beating cilia and mucus production, recapitulating the airway mucosal surface in vivo. Cultures were inoculated with SARS-CoV-2 on the apical surface, washed after 1 hr, then incubated at 35°C to simulate the temperature of the upper respiratory tract and conducting airways. Cultures were collected for RNA isolation and RT-qPCR at 1 hr (post-inoculation time point), 24hr, 48hr, 72hr, and 96hr and basolateral media was collected for CXCL10 ELISA. Reminiscent of what we observed during SARS-CoV-2 infection in vivo (Fig 3D-I), viral load increased rapidly for the first three days of infection, then plateaued between 72-96 hr (Fig 4A). ISGs were also induced, with ISG mRNA levels and CXCL10 protein level in the basolateral medium increasing markedly from 72 to 96 hr (Fig 4B-F). Notably, a very high level of CXCL10 protein was produced by infected epithelia (∼4ng/ml by 96 hr), consistent with the strong NP CXCL10 signal observed in the nasopharynx in vivo.

**Figure 4.**
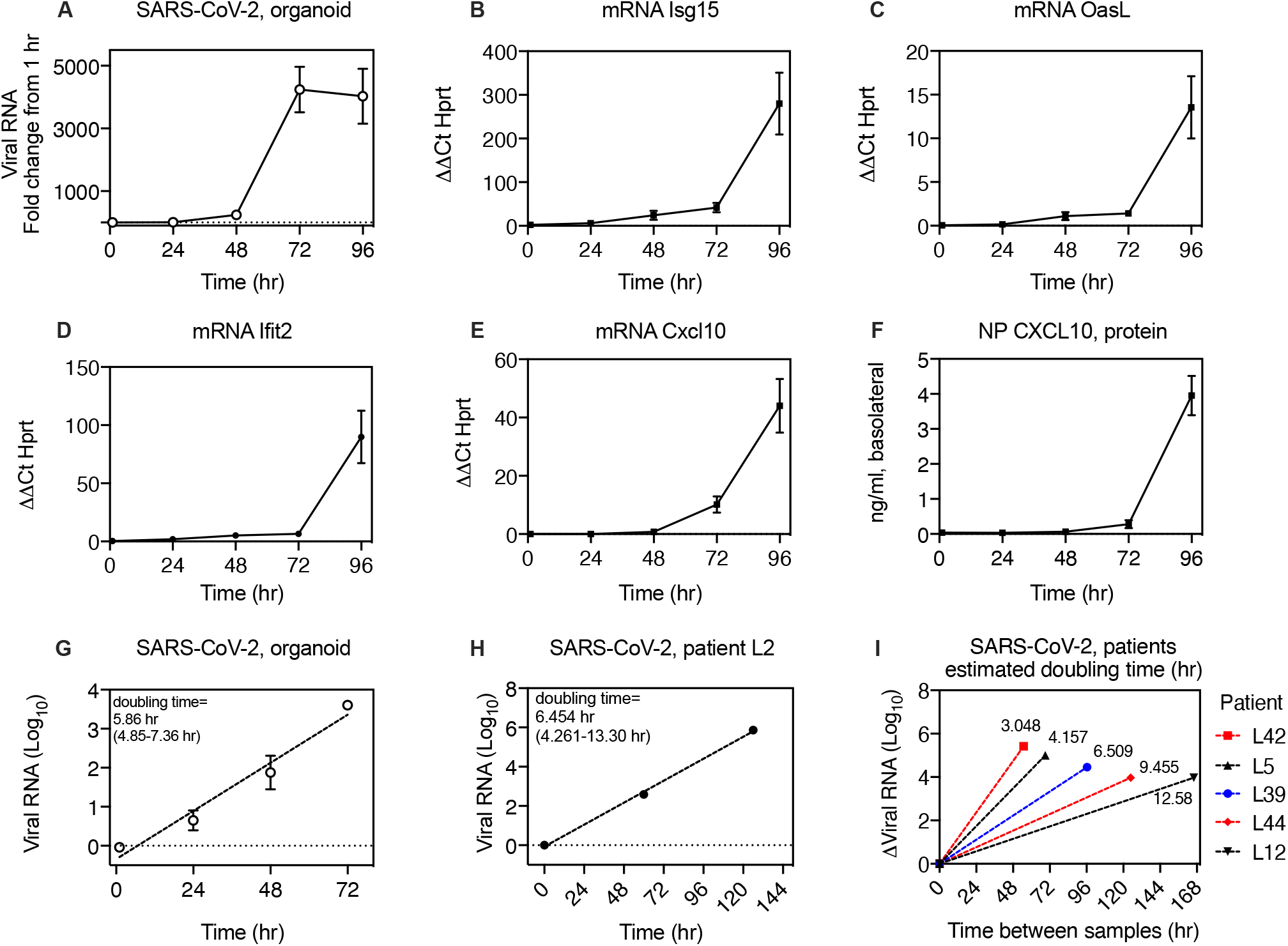
Kinetics of SARS-CoV-2 replication in organoids and in vivo. (A) Time course of SARS-CoV-2 replication in human primary airway epithelial organoids, expressed as fold increase from 1hr (post-inoculation time point). (B) - (E) ISG mRNA level relative to HPRT mRNA in organoids during SARS-CoV-2 infection. (F) CXCL10 protein in the basolateral medium during SARS-COV-2 replication. For A-H, symbols show mean and S.E.M. of five biological replicates per condition. (G) Exponential curve fit for increase in RNA during SARS-CoV-2 during replication in organoids from 1-72 hr and calculated doubling time for exponential growth. (H) Exponential curve fit for increase in viral RNA during SARS-CoV-2 replication for first three virus-positive samples from patient L2 and calculated doubling time for exponential growth with 95% confidence interval. (I) Estimated doubling times for increase in viral RNA during SARS-CoV-2 replication in patients with one SARS-CoV-2 positive sample prior to peak viral load, shown in fig 3 B-F. Y-axis shows change in viral RNA and x-axis shows time interval between samples. Doubling time calculation assumes exponential growth between first and peak viral load samples.

### Doubling time of SARS-CoV-2 in vitro and in vivo

Viral replication in organoid cultures appeared to follow an exponential curve for the first 72hr of infection. Therefore, we used curve-fitting to exponential growth to estimate the doubling time, which was 5.858 hr (95% C.I. of 4.85-7.357 hr., based on 20 y-values, 5 per time point; Fig 4G). For patient L2, viral load data from first three SARS-CoV-2+ time points also appeared to follow exponential growth, therefore we used the same method to estimate the SARS-CoV-2 doubling time in vivo from this data, which was 6.454 hr (95% C.I. 4.261-13.30 hr based on 3 y-values, Fig 4H). For all other patients from whom viral load increased in serial samples (Fig 3), we had only one sample prior to peak viral load. We asked what the doubling times for SARS-CoV-2 would be in these samples if we assumed exponential replication between the first and peak viral RNA values. The calculated doubling times across patients ranged from 3.048-6.509 hr for samples less than or equal to 5 days apart. For the two patients with a larger sampling interval (L44, L12), calculated doubling times were 9.455 and 12.58 hr, although these calculations would be expected to overestimate doubling time if the second sample was taken after viral replication had plateaued or begun to decline. Together, the in vitro and in vivo results indicate that SARS-CoV-2 replicates exponentially during the first few days post-infection prior to the peak host anti-viral response, with an average doubling time of approximately 6 hr.

### Effect of prior rhinovirus infection on ISG induction and SARS-CoV-2 replication

One of the many physiological exposures that could potentially alter the local innate immune response to SARS-CoV-2 in the upper airway mucosa is recent infection by other viruses. To model this situation, we used organoid culture to examine the effects of prior exposure to rhinovirus, the most frequently detected virus in the human upper respiratory tract, on subsequent SARS-CoV-2 infection (Fig 5A). Based on previous studies, we expected that rhinovirus might curtail infection by inducing an epithelial antiviral response, but also could potentially promote infection by increasing expression of SARS-CoV-2 entry receptors (Wu et al., 2020; Ziegler et al., 2020). Similar to previous observations, rhinovirus infection (HRV-01A, MOI ∼0.05) led to robust induction of interferon stimulated genes by day 3 post-infection (Fig 5B). We then evaluated whether rhinovirus infection altered the expression of ACE2, the SARS-CoV-2 entry receptor. ACE2 was originally reported to be an ISG, but a subsequent study reported that full-length ACE2, which functions as an entry receptor, is not an ISG, and that a truncated form, dACE2, is an ISG but is not a functional SARS-CoV-2 entry receptor (Onabajo et al., 2020; Ziegler et al., 2020). Consistent with this finding, we observed that dACE-2 was significantly induced by rhinovirus infection (∼14-fold) and that, as expected for an ISG, induction was prevented by blocking activation of IRF3, a transcription factor downstream of viral RNA sensors, using the inhibitor BX795 (Clark et al., 2009). In contrast, full-length ACE2 expression was slightly but significantly increased by rhinovirus infection(∼2-fold) and this change was not abrogated by BX795, suggesting a different mechanism of induction (Fig S3.) Rhinovirus infection had no effect on expression of TMPRSS2 (not shown).

**Figure 5.**
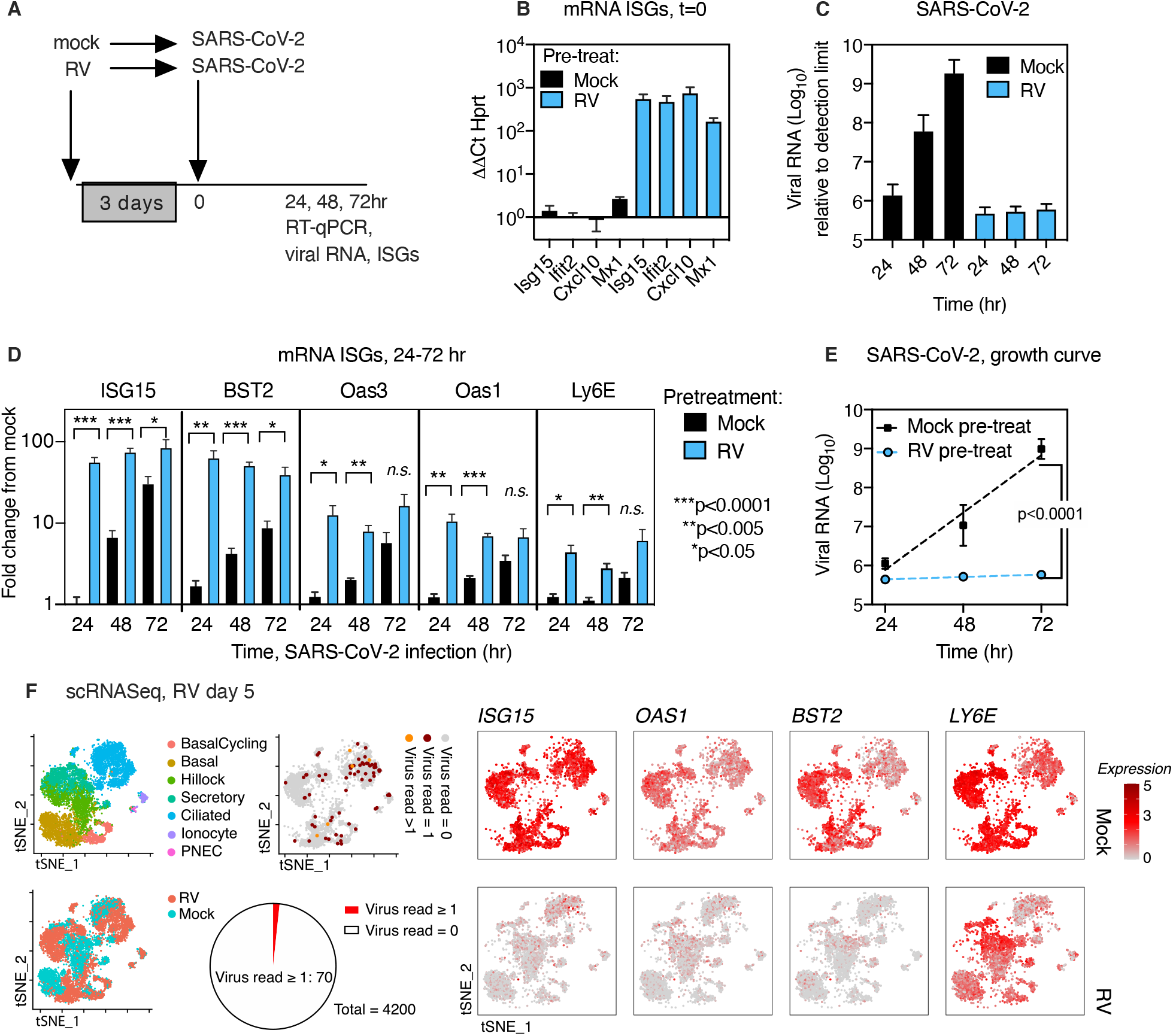
Effect of prior rhinovirus infection on ISG induction and SARS-CoV-2 replication in human airway epithelial organoids. (A) Timing of infection of epithelial organoids with rhinovirus followed by SARS-CoV-2 (B) Expression of interferon stimulated genes in airway epithelial organoids 3 days post-rhinovirus infection, relative to mRNA for the housekeeping gene HPRT (C) SARS-CoV-2 viral RNA at 24, 48, and 72 hr post-infection, with or without RV pre-infection (D) Expression of interferon stimulated genes at 24, 48, and 72 hr post SARS-CoV-2 infection, with or without RV pre-infection, expressed as fold change from uninfected cells (E) Replication of SARS-CoV-2 in mock-vs. rhinovirus-pretreated cultures, fit to exponential growth curve (F) Single cell sequencing of human airway epithelial cell organoids, mock or 5 days post rhinovirus infection. Red and orange dots indicate 70/4200 cells with detectable viral RNA at this time point in rhinovirus-infected cultures (at least 1 read from viral RNA). TSNE plots show expression of mRNA for ISGs in mock and infected cultures at the same time point.

Next, we evaluated SARS-CoV-2 replication and ISG induction following infection of airway epithelial organoids, with or without prior rhinovirus infection. SARS-CoV-2 viral load increased exponentially in infected cultures without prior RV infection, as observed previously (Fig 4), but showed essentially no increase when cultures had been exposed to rhinovirus 3 days prior (Fig 5C). Evaluation of ISG expression over the course of infection showed that at early time points of SARS-CoV-2 infection (24, 48 hr, and sometimes 72hr), ISGs were significantly more highly expressed in RV-preinfected cultures that in cultures infected with SARS-CoV-2 without prior RV exposure (Fig 5D). This included several ISGs which have been previously reported to limit coronavirus replication or for which polymorphisms are linked to disease severity of SARS-CoV or SARS-CoV-2, including ISG15, BST2 (tetherin), and LY6E, and OAS1-3 (Hamano et al., 2005; He et al., 2006; Ma et al., 2014; Martin-Sancho et al., 2020; Pairo-Castineira et al., 2020; Pfaender et al., 2020; Taylor et al., 2015) (Fig 5D). Non-linear regression analysis was consistent with exponential replication of SARS-CoV-2 from 24-72 hr post infection in mock-pretreated cultures, in contrast to rhinovirus pre-infected cultures which supported essentially no replication (Fig 5E).

To better understand the timing and breadth of the epithelial host response to rhinovirus that appeared to limit SARS-CoV-2 replication, we evaluated ISG expression over time for five days post rhinovirus infection and examined ISG expression and viral infection at the single cell level. Time course analysis showed that following inoculation, rhinovirus replicated robustly, peaking at 24 hr post-infection, and then declined significantly but was still detectable by RT-qPCR at day 5, a time point corresponding to 48 hr post SARS-CoV-2 infection in the sequential infection experiment (FigS4A). ISG expression increased and decreased in parallel with viral replication but was still significantly higher than in mock-treated cells at day 5 post-rhinovirus infection(Fig S4B-E). Next, we performed single cell RNA sequencing to evaluate the host ISG response in infected and bystander cells on day 5 post rhinovirus infection. At this time point, rhinovirus viral RNA reads were detected in only 70 out of 4200 cells sequenced (Fig 5F). The infected cells were predominantly ciliated cells but included all major cell types, consistent with the HRV-01A entry receptor LDL-R being ubiquitously highly expressed throughout the culture (Fig S4F,G). Although rhinovirus was only detected in a small subset of cells in infected cultures at day 5 (1.67%), ISGs were elevated in all cells compared to mock-treated cultures (Fig 5F), demonstrating that rhinovirus infection induces a robust bystander antiviral response in uninfected cells that lasts at least 5 days.

### Blocking ISG induction restores SARS-CoV-2 replication following rhinovirus infection

Next, to test whether suppression of SARS-CoV-2 replication by rhinovirus was dependent upon the host cell interferon response, we pre-treated cells with the signaling inhibitor BX795 18hr before rhinovirus infection, which prevents interferon and ISG induction by rhinovirus(Fig 6A, B)(Clark et al., 2009; Wu et al., 2020). There was no significant difference in SARS-CoV-2 viral load in RNA isolated from organoid cultures at 72 hr for cultures with and without drug treatment, (Fig 6B). The effect of BX795 treatment was much more striking in the setting of sequential rhinovirus-SARS-CoV-2 infection. As seen previously (Fig 5), prior infection with rhinovirus suppressed SARS-CoV-2 replication by >1000-fold, but replication was restored by BX795 pre-treatment (Fig 6C). These results indicate that IRF3 signaling is critical for the inhibition of SARS-CoV-2 replication by prior rhinovirus infection, consistent with the effects of RV pre-infection on ISG induction (Fig 5).

**Figure 6.**
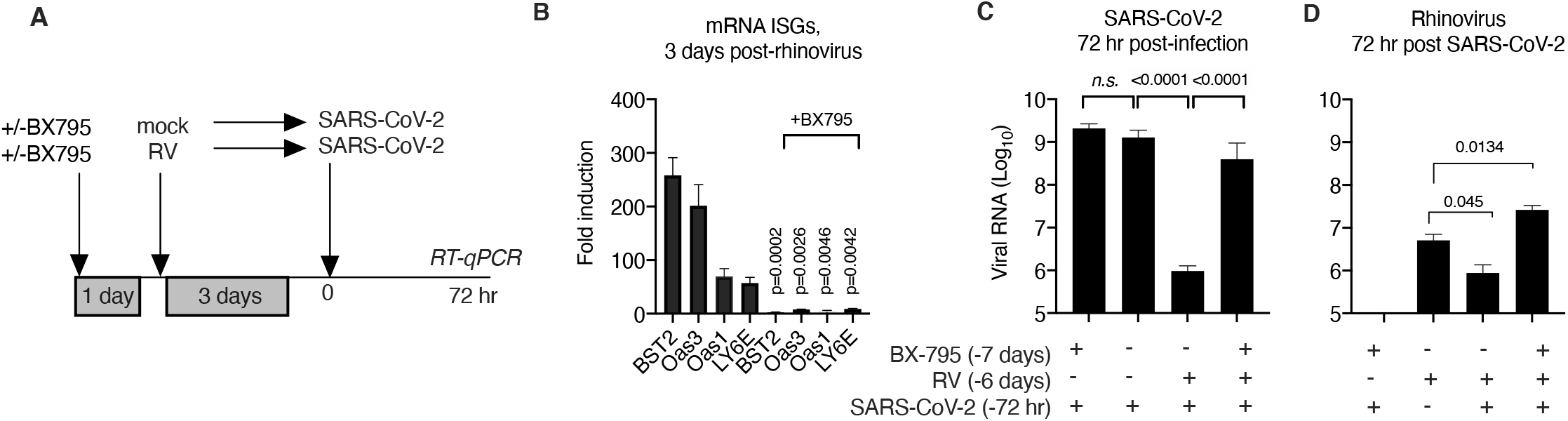
Effect of pretreatment with BX795 during sequential rhinovirus, SARS-CoV-2 infection. Organoid cultures were pretreated with or without BX795 for 18 hr, then mock-infected or infected with HRV01A, incubated for 3 days, then infected with SARS-CoV-2. (A) Effect of BX-795 pre-treatment on ISG induction, 3 days post rhinovirus infection. Bars show fold change in ISG mRNA level in RV infected cultures compared to mock without (left) or with (right) BX-795 pre-treatment. P values indicate significant differences between ISG levels in cultures with or without BX795 pretreatment by t-test. (B) SARS-CoV-2 viral RNA level relative to the limit of detection in organoid cultures, 72 hr post SARS-CoV-2 infection, with and without BX-795 and/or RV pre-treatment. P values indicate significant differences in viral RNA levels, n.s. = not significant. (C) HRV01A viral RNA level relative to the limit of detection in organoid cultures, 72 hr post SARS-CoV-2 infection, with and without BX-795 and/or RV pre-treatment. This graph also includes cultures infected with RV but not subsequently infected with SARS-CoV-2. P values indicate significant differences in viral RNA levels. For all graphs, bars show mean and S.E.M. of 4-6 biological replicates per condition.

RV viral RNA was detected at much lower levels than SARS-CoV2 RNA at this time point (72hr post SARS-CoV-2 infection, 6 days post-RV infection), and showed a slight reduction during SARS-CoV-2 coinfection without BX-795, but significantly higher levels during co-infection in the presence of BX-795. This result indicates that the antiviral response limits rhinovirus replication in the setting of SARS-CoV-2 co-infection at this time point, and that both viruses achieve higher viral loads with inhibition of innate antiviral signaling. In other words, in the presence of an intact antiviral response, viral load of both viruses is reduced by co-infection, but if the host response is inhibited, the viral load of both viruses is higher during co-infection. This experiment models viral co-infection in a host with an intact interferon response (both viruses decrease during co-infection), compared to a host with a deficient interferon response (equal or greater replication of both viruses during co-infection.)

### Blocking ISG induction enhances SARS-CoV-2 replication in a low MOI infection

Next, we further probed the effect of BX-795 treatment on SARS-CoV-2 replication using a ten-fold lower MOI (MOI ∼0.05), conditions under which SARS-CoV-2 would potentially be more sensitive to suppression by ISGs. Under these conditions, BX795 treatment led to a ∼10-fold increase in intracellular viral load and ∼300-fold increase in virus shedding into the apical wash at 72 hr post infection. A trend towards an increase (5-10x) was also seen at 96 hrs post-infection, although due to variability among replicates this difference was not statistically significant (Fig 7A, B). Based on these results, we checked the effect of BX795 on viral shedding into the apical wash in the MOI 0.5 infection (Fig 6) but in this case results were similar to those observed in organoid cultures (not shown). Next, based on the increase in viral RNA from 1hr to 72 hr during the low MOI infection, we estimated the effect of BX795 on the SARS-CoV-2 doubling time in organoid culture. Assuming exponential growth between 1hr and 72 hr, the SARS-CoV-2 doubling time without BX795 was 5.127 hr (95%C.I. 3.889 to 7.518 hrs), and with BX795 was 3.578 hr (95%C.I. 3.499 to 3.661). We then examined expression of ISGs that have been shown to limit coronavirus replication and which have high basal expression in airway epithelial cultures including IFITM3, ISG15, and BST2 (tetherin), thus could be particularly important for limiting a low MOI infection. Induction of all of these ISGs was suppressed by BX795 pre-treatment (Fig 7D-F). For BST2 mRNA, blocking ISG induction with BX795 revealed a decrease in BST2 mRNA below the baseline level during SARS-CoV-2 infection, suggesting that SARS-CoV-2 may antagonize BST2 expression at the mRNA level, in addition to other mechanisms whereby SARS-CoV-2 or SARS-CoV have been reported to antagonize BST2 at the protein level (Martin-Sancho et al., 2020; Taylor et al., 2015). The effects of BX795 on SARS-CoV-2 during low MOI infection indicate that the epithelial antiviral response induced by SARS-CoV-2 does limit viral replication, albeit to a lesser extent than interference by rhinovirus.

**Fig 7.**
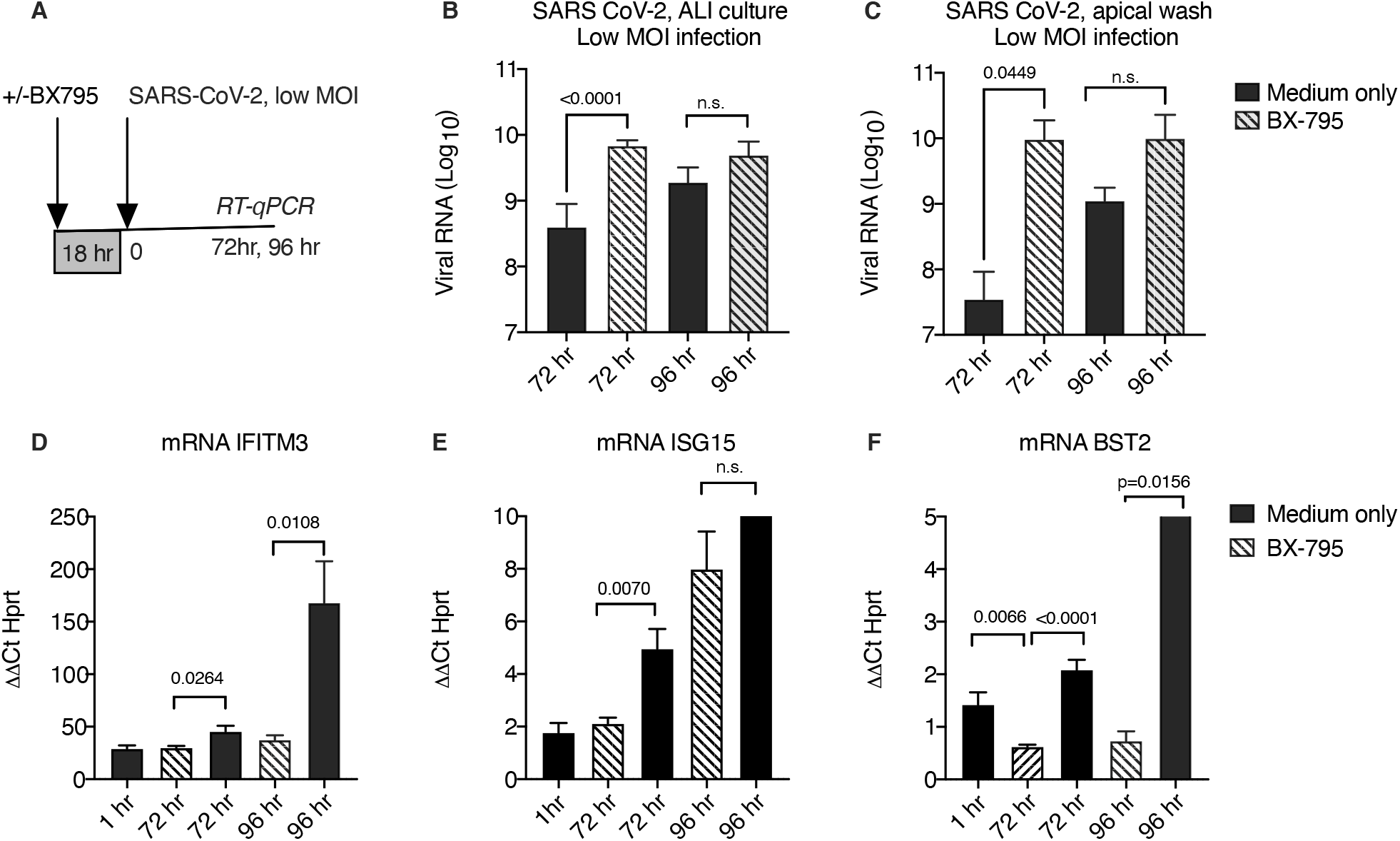
Effect of BX795 on SARS-CoV-2 replication in low MOI infection. (A) Cultures were pre-treated with 6μM BX-795 or medium only, then inoculated with SARS-CoV-2, MOI 0.05, at t=0 hr. Cultures were collected for RNA isolation and RT-qPCR at t=1, 72, and 96 hr post infection. Apical wash was collected at 72 and 92 hr post-infection. (B)SARS-CoV-2 viral RNA in RNA isolated from organoid cultures 72 hr and 96 hr post-inoculation, with (hatched bars) or without BX795 (black bars), relative to the limit of detection. (C)SARS-CoV-2 viral RNA in RNA isolated from the apical wash of cultures collected at 72 hr and 96 hr post-inoculation, with (hatched bars) or without BX795 (black bars), relative to the limit of detection. (D)-(F) ISG mRNA level in SARS-CoV-2 infected cultures, graphed relative to the mRNA level for the housekeeping gene HPRT (2^Ct-Ct^ ^HPRT^), at 1, 72, or 96 hr post-inoculation. (B)-(F) Bars show mean and S.E.M. for 4-6 biological replicates per condition. P-values are shown for significant differences, based on the students t-test.

Taken together, our results show that SARS-CoV-2 undergoes exponential replication at the start of infection and induces ISG expression in vitro and in vivo, and that conditions which alter the initial ISG expression level or rate of ISG induction, including prior infection by a different virus, can profoundly impact SARS-CoV-2 replication.

## Discussion

SARS-CoV-2 replication in the nasopharynx is known to peak during the first week of infection, but the biological variables governing the rate and magnitude of viral amplification are not fully understood (Cevik, 2020; He et al., 2020). Nasopharyngeal viral load early in infection correlates with likelihood of transmitting the infection, and robust viral amplification in the respiratory tract is also likely a prerequisite, although certainly not the only factor, for progression of COVID-19 disease. Here we present evidence that during the first few days of SARS-CoV-2 infection, the airway interferon response plays a protective role by curtailing viral replication in its initial target tissue: the airway epithelium. Specifically, our data show that the extent of viral replication is determined by the magnitude and timing of the host interferon response during a critical time window at the start of infection.

The interferon response is a potent mechanism of antiviral innate defense at mucosal surfaces and effectively curtails replication of many viruses, most of which also antagonize this host response to some degree in order to enable viral replication(Garcia-Sastre, 2017; Iwasaki, 2012). Viral recognition by innate immune sensors within infected epithelial cells induces expression of type I and type III interferons and interferon stimulated genes (ISGs), a diverse family of antiviral effectors, both directly, in infected cells, and in neighboring cells through paracrine effects of secreted interferons(Odendall and Kagan, 2015; Schneider et al., 2014). SARS-CoV-2 has been reported to induce interferons and ISGs in the airway mucosa in vivo and in vitro (Mick et al., 2020; Mulay et al., 2020; Ravindra et al., 2020; Zhou et al., 2020). Consistently, we observed robust ISG induction in all patients using RNA-Seq of nasopharyngeal RNA, regardless of disease severity or other biological variables (Fig 1). However, there is also convincing evidence that SARS-CoV-2 antagonizes the interferon response in infected cells through multiple mechanisms(Banerjee et al., 2020; Konno et al., 2020; Martin-Sancho et al., 2020; Xia et al., 2020). Our observations with organoid culture (Fig 4), and in the one patient for whom we had multiple longitudinal samples prior to peak viral load (patient L2, Fig 3A) show exponential viral replication at the start of SARS-CoV-2 infection. ISGs are also induced, but it is unclear when this host response becomes functionally effective. We found that in the first 72 hours of infection, prior to the peak host response, blocking ISG induction had no effect in SARS-CoV infection at MOI 0.5 (Fig 6), but did increase viral replication at ten-fold lower MOI (Fig 7; MOI 0.05). Together, these observations support a model in which antagonism by SARS-CoV-2 attenuates but does not prevent the interferon response and thereby creates a time window at the start of infection during which the virus can undergo exponential growth.

Due to the nature of exponential growth, a small change in biological variables affecting either the rate of viral replication or the rate of development of an effective interferon response could have a profound impact on viral amplification and peak viral load. For example, based on an average viral doubling time estimated from our data of ∼6 hr, a 24 hour delay in the development of an effective interferon response, for example due to host deficiency in innate immune signaling, would lead to a 16-fold increase in peak viral load. Thus, a peak viral load of 1000 infectious particles would become 16,000 infectious particles, an upper airway viral load much more likely to lead to viral transmission or spread of the infection to the lower respiratory tract. Likewise, if conditions or viral differences allowed the viral doubling time to decrease by two hrs, from 6 hr to 4hr, this would lead to 16-fold greater viral amplification by 48 hr, and 64-fold greater amplification by 72hr, for the faster growing strain. The extent to which NP viral load correlates with disease severity is still unclear, although several studies show an association (Huang et al., 2020; Liu et al., 2020; Yilmaz et al., 2020; Zheng et al., 2020). However, there is strong evidence that high NP viral load correlates with viral transmission, an issue that has come into focus recently due to the emergence of SARS-CoV-2 strains that appear to have enhanced transmission, such as the B.1.1.7 strain emerging in the U.K.(Cevik, 2020; He et al., 2020; Prevention, 2021; Singanayagam et al., 2020) Our study predicts biological features of the virus that could potentially underlie this greater transmissibility, such as better antagonism of the host interferon response or faster doubling time, both of which would be expected to increase NP viral load and therefore increase transmissibility.

Conversely, host factors that enhance interferon-mediated defenses could be impactful in reducing peak viral load or even preventing infection altogether. One factor that is known to alter ISG expression in the nasopharynx is recent viral infection. In this study, we focused on rhinovirus, the most frequent cause of the common cold. Recent epidemiological studies have shown that this virus is much more prevalent in the upper respiratory tract than previously appreciated (Foxman and Iwasaki, 2011; Jartti et al., 2008). For example, in a recent year-round study, about one-third (34%) of all nasal samples from young children (<5 yrs) were rhinovirus-positive, regardless of symptoms (Byington et al., 2015). Rhinoviruses and other respiratory viruses have also been shown to induce ISGs in the nasopharynx in vivo, in both the presence and absence of symptoms (Landry and Foxman, 2018; Wolsk et al., 2016; Yahya et al., 2017; Yu et al., 2019). Thus, rhinovirus fits the characteristics of a common environmental factor that could impact ISG expression in the respiratory tract.

Our data using sequential infection in an airway epithelial organoid model show that prior rhinovirus infection blocks replication of SARS-CoV-2 that this protection is dependent upon ISG induction. Furthermore, using time course studies and single cell analysis of organoid cultures, we found that a significant bystander interferon response can be detected in cells throughout the epithelium for at least 5 days post-rhinovirus infection, even though viral replication peaks at 24 hr and few infected cells are detected at this time point (Fig 5, Fig S4). This finding is consistent with recent work from our group and others showing that the host interferon response triggered by one respiratory virus can block infection by another (Essaidi-Laziosi et al., 2020; Wu et al., 2020). Viral interference seen during sequential influenza virus infections was in fact the basis for the original discovery and naming of interferons in 1957, but only recently has this idea been explored in depth with regards to human respiratory viral infections, triggered in part by epidemiological data suggesting interference among RNA respiratory viruses(Greer et al., 2009; Isaacs and Lindenmann, 1957; Karppinen et al., 2016; Nickbakhsh et al., 2019; Schultz-Cherry, 2015). ISG induction by an unrelated virus may be particularly effective against a virus like SARS-CoV-2, since it would pre-empt many of the mechanisms SARS-CoV-2 has in place to antagonize interferon and ISG induction in response to its own replication, including specific targeting of ISGs effective in blocking the coronavirus life cycle such as BST2 (Martin-Sancho et al., 2020; Taylor et al., 2015).

The concept of viral interference based on the host interferon response assumes a host with intact innate immune defenses, which may not always be the case. In our experimental model, there was a profound difference in the outcome of rhinovirus-SARS-CoV-2 co-infection in the presence and absence of an intact host cell interferon response (Fig 6). With an intact host response, viral loads of both viruses decreased, but when the interferon response was blocked, viral loads of both viruses were equal to or higher than in single infections (Fig 6.) This result illustrates that the expected outcome of a viral co-infection is not one-size-fits-all: it is likely to be profoundly dependent upon host innate immune status. Many host factors ranging from genetic polymorphisms to transient environmental conditions in the airway (e.g. temperature, humidity) can attenuate interferon responses and promote replication of respiratory viruses, and would likewise be expected to reduce the interferon-mediated protective effects of viral co-infection (Asgari et al., 2017; Foxman et al., 2015; Kudo et al., 2019; Lamborn et al., 2017; Mihaylova et al., 2018; Zhang et al., 2020). Also, interference requires closely spaced virus co-exposures, which may be less frequent at certain times of the year, or during the use of pandemic mitigation measures. Low rates of influenza during 2020 in the southern hemisphere indicate that the public health measures put in place to slow the spread of SARS-CoV-2 also suppressed circulation of other respiratory viruses, although there is some evidence that rhinoviruses have continued to circulate(Olsen et al., 2020; Poole et al., 2020). Viral interference may become a more important consideration for understanding susceptibly to COVID-19 and other viral pathogens as society reopens.

There are several important caveats to our study. First, our data indicate a relatively limited effect of the SARS-CoV-2-induced interferon response on initial viral replication when it is the only virus present, at least at high MOI (Fig 6). However, the recruitment of cells of the immune system to the respiratory tract, as indicated by our RNAseq data (Fig 1), could considerably amplify ISG induction in vivo. Recent work showing that interferon deficiencies are linked to severe COVID-19 indicates the importance of interferon-mediated defense against this virus in vivo (Bastard et al., 2020; Pairo-Castineira et al., 2020; Zhang et al., 2020). Second, in vivo virus-host-virus interactions could be more complicated than those seen in organoid culture; for example, if a viral infection results in residual lung damage this could exacerbate rather than protect against a subsequent viral respiratory illness. Studying virus-host-virus interactions in vivo will be an important future direction of this study. For assessing viral interference, it will be critical not only to look at co-occurrence or sequential occurrence of viral infections, but also to measure the antiviral response in vivo. For example, symptomatic upper respiratory tract infections induce greater ISG responses than asymptomatic, and the latter may not cause significant interference(Yahya et al., 2017). Such studies will be facilitated by increasing development of nasopharyngeal swab-based methods to assess the airway host immune response, as presented here and in the recent literature(Landry and Foxman, 2018; Mick et al., 2020; Wolsk et al., 2016; Yu et al., 2019).

In sum, our results demonstrate an important role for the interferon-mediated defenses in curtailing SARS-CoV-2 replication at the start of infection and compel further studies of how the changing conditions present in the upper respiratory tract, including recent infection by other viruses, can modulate host antiviral defenses and alter SARS-CoV-2 infection and transmission.

## Supporting information

Cheemarla et al_2021_Supplemental Figures and Table

## Data Availability

We plan to deposit RNASeq data sets in the GEO database and this data will be publicly available at the time of study publication in a peer-reviewed journal.

## Acknowledgements

We would like to thank Craig Wilen and Wilen lab members for valuable help and advice, and for providing SARS-CoV-2 virus stock. We thank Maureen Owen, Robin Garner, Greta Edelman, the entire staff of the Yale New Haven Hospital Clinical Virology laboratory, and Amy Likens for their dedicated assistance. We also thank Bryan Pasqualucci and Christopher Castaldi at the Yale Center for Genomic Analysis. Funding was provided by Fast Grants (Emergent Ventures, Mercatus Institute, George Mason University) to E.F.F., the Yale Department of Laboratory Medicine and COVID-19 Dean’s Fund (E.F.F.), the China Scholarship Council-Yale World Scholars Fellowship (B.W.), the Gruber Foundation Fellowship (T.W.), and the NIH (T32AI007019, T.W.).

## Author Contributions

Conceptualization E.F.F.; Methodology, N.R.C., T.W., V.T.M., B.W., D.Z., M.L.L., and E.F.F.; Investigation N.R.C., T.W., V.T.M., B.W., and E.F.F.; Formal analysis B.W., D.Z., and G.W.; Resources, M.L.L.; Writing – Original Draft, E.F.F.; Writing – Review & Editing, N.R.C., T.W., V.T.M., B.W.,D.Z., G.W., M.L.L., E.F.F.; Funding Acquisition, E.F.F.

## Methods details

### RESOURCE AVAILABILITY

#### Lead Contact

Further information and requests may be directed to, and will be fulfilled by the lead contact Ellen F. Foxman

#### Materials availability

This study did not generate new reagents.

#### Data and Code Availability

Nasopharyngeal transcriptome data and single cell RNA sequencing data will be publicly available in the GEO database at the time of publication.

### EXPERIMENTAL MODEL AND SUBJECT DETAILS

#### Ethics statement

The use of clinical samples and data in this study was approved by the Yale Human Research Protection Program Institutional Review Board (Protocol ID #200002765). Procedures for testing residual clinical samples and recording linked patient data, followed by sample and data de-identification, were evaluated and the requirement for specific patient consent was waived. In vitro experiments used primary human cells obtained from Lonza Bioscience (Walkersville, Maryland, U.S.A.). Lonza guarantees that all tissue utilized for human cell products is ethically obtained with donor informed consent in accordance with processes approved by an Institutional Review Board or comparable independent review body.

#### Clinical samples

We used viral residual nasopharyngeal (NP) samples remaining after clinical testing for CXCL10 measurements and transcriptome analysis. Swab-associated viral transport medium was stored at -80 °C following clinical testing and thawed just prior to ELISA assay or RNA isolation for RNA-Seq. Clinical information including age, sex, virology results, and specific features of clinical course including presenting symptoms, hospital admission and length of stay, was extracted from the electronic medical record and recorded, after which samples were assigned a study code and de-identified. In the clinical laboratory, SARS-CoV-2 was detected in most samples using an EUA-approved TaqMan assay detecting the CDC targets N1, N2, and RNAseP (Prevention, 2020). In some longitudinal samples, SARS-CoV-2 was diagnosed with the commercial Cepheid assay (reference); in this case, RT-qPCR for the CDC N1 gene was repeated using RT-qPCR TaqMan assay for the CDC N1 gene as described previously (Cat no: 10006600, Integrated DNA Technologies, IA)(Vogels et al., 2020).

#### Primary human bronchial epithelial cells

Primary human bronchial epithelial cells from healthy adult donors were obtained commercially (Lonza, Walkersville, MD, USA) and cultured at air-liquid interface according to manufacturer’s instructions using reduced hydrocortisone (Stem Cell Technologies, Vancouver, Canada). Cells were allowed to differentiate for four weeks by which time they displayed beating cilia and mucus production.

#### Viruses

Rhinovirus 1A (HRV-01A; ATCC VR-481) was amplified in H1-HeLa cells (ATCC CRL-1985) and titer was determined by plaque assay as reported previously (Foxman et al., 2015). SARS-CoV-2 (BEI resources, USA-WA1/2020) was generously provided by the Wilen lab. Virus was cultured on Vero E6 cells and titer was determined by plaque assay as described previously (Ravindra et al., 2020).

#### RNA isolation from clinical samples

At the time of accessioning, the residual viral transport medium from clinical samples was stored at -80C. Upon thawing, RNA was isolated from 140μl of transport medium using the Qiagen Viral RNA isolation kit per manufacturer’s instructions (Ref: 52904, Qiagen, Germany) and one aliquot was reserved for ELISA.

#### Library preparation and RNA Sequencing

RNA samples were quantified and checked for quality using the Agilent 2100 Bioanalyzer Pico RNA Assay. Library preparation was performed using Kapa Biosystem’s KAPA HyperPrep Kit with RiboErase (HMR) in which samples were normalized with a total RNA input of 25ng. Libraries were amplified using 15 PCR cycles. Libraries were validated using Agilent TapeStation 4200 D1000 assay and quantified using the KAPA Library Quantification Kit for Illumina® Platforms kit. Libraries were diluted to 1.3nM and pooled at 1.25% each of an Illumina NovaSeq 6000 S4 flowcell using the XP workflow to generate 25M read pairs / sample.

#### RNA-Seq data analysis

Low quality reads were trimmed and adaptor contamination was removed using Trim Galore (v0.5.0, https://www.bioinformatics.babraham.ac.uk/projects/trim_galore/). Trimmed reads were mapped to the human reference genome (hg38) using HISAT2 (v2.1.0)(Kim et al., 2019). Gene expression levels were quantified using StringTie (v1.3.3b) with gene models (v27) from the GENCODE project(Pertea et al., 2015). Differentially expressed genes (adjusted p value < 0.05, fold change cutoff = 2) were identified using DESeq2 (v 1.22.1) (Love et al., 2014). To avoid the unexpected outlier replacement for sex-linked genes, we turned off the outlier replacement option in the male vs. female comparison by setting minReplicatesForReplace=Inf for the DESeq() function in the DESeq2 package.

#### Visualization of RNA-Seq data

Protein coding genes differentially expressed in SARS-CoV-2+ vs. negative control were visualized on a volcano plot, with an x-axis cutoff l log_2_FC cutoff l =10. All differentially expressed RNAs are included in Table S1 (n=1770). Significantly differentially expressed transcripts were defined as those with log_2_FC>1 and adjusted p value<0.05. Heatmap shows gene expression levels of top 45 most significant DEGs using min-to-max scaling of normalized read counts.

Pathway analysis was performed using Ingenuity Pathway Analysis (version 01-16). Transcription factor motif enrichment analysis was performed using Cytoscape (version 3.8.1) with the iRegulon plug-in ((version 1.3)(Janky et al., 2014).

#### In vitro infections

We infected primary human bronchial epithelial cells differentiated at air-liquid interface with HRV-01A, SARS-CoV-2, or both. For SARS-CoV-2, high MOI infection was MOI 0.5 and low MOI infection was MOI 0.05. For HRV-01A, MOI 0.1 was used, as this was the minimum viral inoculum that reproducibly led to robust HRV-01A viral replication in ALI cultures based on prior studies.

To evaluate the effect of RV on subsequent infection with SARS-CoV-2, we infected with each virus individually or sequentially and examined the time course of viral amplification and ISG induction. To formally test whether prior exposure to RV inhibits SARS-CoV-2 replication through activation of the host cell interferon response, we performed sequential infection studies in the presence of BX795.

#### RT-qPCR

For RT-qPCR, RNA was isolated from each well of differentiated epithelial cells using the QIAGEN RNeasy kit by incubating each 24-well insert with 350 μl lysis buffer at room temperature for 5 minutes, followed by RNA isolation and cDNA synthesis using iScript cDNA synthesis kit (BioRad). To quantify viral RNA and mRNA levels for interferon stimulated genes and the housekeeping gene HPRT, qPCR was performed using SYBR green iTaq universal (BioRad) per manufacturer’s instructions. Viral RNA was quantified using primers to the RV genome. Viral RNA per ng total RNA is graphed as fold change from the limit of detection (40 cycles of PCR) as 2^40-Ct^. ISG mRNA levels are graphed as fold change from mock-treated cells or are presented relative to the level of mRNA for the housekeeping gene HRPT (2^-ΔΔCT^). RT-qPCR for SARS-CoV-2 within cultures was performed using the previously-described TaqMan assay for the CDC N1 gene with primers and probes provided by IDT (Cat no: 10006600, Integrated DNA Technologies, IA). RT-qPCR for SARS-CoV-2 in apical was performed using a combined reverse transcriptase and qPCR reaction using the Luna Universal Probe One-Step RT-qPCR Kit (New England Biolabs, MA)

The following primers were used for RT-qPCR with SYBR green:

HPRT (F-TGGTCAGGCAGTATAATCCAAAG; R-TTTCAAATCCAACAAAGTCTGGC)

ISG15 (F-CATCTTTGCCAGTACAGGAGC; R-GGGACACCTGGAATTCGTTG)

RSAD2 (F-TCGCTATCTCCTGTGACAGC;R-CACCACCTCCTCAGCTTTTG)

MX1 (F-AGAGAAGGTGAGAAGCTGATCC;R-TTCTTCCAGCTCCTTCTCTCTG)

IFITM3 (F-ATCGTCATCCCAGTGCTGAT;R-ATGGAAGTTGGAGTACGTGG)

IFIT2 (F-CCTCAAAGGGCAAAACGAGG; R-CTGATTTCTGCCTGGTCAGC)

CXCL10 (F-CCTGCAAGCCAATTTTGTCC; R-ATGGCCTTCGATTCTGGATTC

LY6E (F-GCATTGGGAATCTCGTGACA; R-ATGGAAGCCACACCAACATT)

BST2 (F-CACACTGTGATGGCCCTAAT; R-TGTAGTGATCTCTCCCTCAAGC)

IFITM3 (F-ATCGTCATCCCAGTGCTGAT; R-ATGGAAGTTGGAGTACGTGG)

OAS1 (F-GCTCCTACCCTGTGTGTGTGT; R-TGGTGAGAGGACTGAGGAAGA)

OAS3 (F-GAAAACTGTCAAGGGAGGCTC; OAS3 R-CCCTCTGGTCCACATAGCTC)

HRV-01A (F- CAGGCCAAATTAAAGTCAATAAGC; R- AGGCTGAAGTTTGGTTTTGC)

### Quantification and statistical analysis, RT-qPCR data

GraphPad Prism (version 9.0.0) was used for data analysis. Data are shown as mean +/- S.E.M. Statistical significance of differences between conditions was determined by t tests (two-tailed). Linear regression analysis was used to determine association between clinical parameters, such as viral load and NP CXCL10 in clinical samples and to test the null hypothesis that the slope of the association was significantly different from zero. Non-linear regression analysis was used to fit viral growth to an exponential curve (exponential growth with log(population)) to determine virus doubling times and to test the null hypothesis that one curve fit both data sets for SARS-CoV-2 growth curve with and without rhinovirus pre-infection. p<0.05 was considered statistically significant.

### ELISA

CXCL10 levels in cell-free residual nasopharyngeal swab samples or tissue culture supernatants was quantified using a solid phase sandwich enzyme linked immunosorbent assay (ELISA) (Cat no: DY266, R&D systems, MN). Briefly, frozen viral transport medium from residual nasopharyngeal swab samples or basolateral medium from organoid cultures was thawed on ice and centrifuged to remove cell debris. ELISA was performed according to the manufacturer’s instructions.

### Single cell RNA-Seq of ALI organoid cultures

#### Library preparation and sequencing

Organoid cultures were digested with trypsin/EDTA to form a single cell suspension. The 10X genomics Single Cell 3’ Protocol was used to produce Illumina-ready sequencing libraries with standard Illumina paired-end constructs, according to the manufacturer’s instructions.

#### Analysis of scRNA-Seq data

All downstream analyses were implemented using R version 3.6.3 and the package Seurat v3.1.4(Stuart et al., 2019). The gene expression matrix mock and RV1A infected sample were first individually analyzed in this procedure: Genes expressed in less than 3 cells and cells expressing less than 200 genes were discarded. A distribution histogram of UMI count in all cells was made and cells with less than 8000 UMI counts were discarded. This resulted in a matrix of 21086 genes expressing in 4511 cells in mock sample, and a matrix of 21195 genes expressing in 4200 cells in RV1A infected sample. The raw counts were normalized using the Seurat function NormalizeData with normalization.method = “LogNormalize” and scale.facto = 10000. All genes are scaled using Seurat function ScaleData with default parameters.

Variable features were determined using method “vst”. A total of top 2000 variable features were used for principal component analysis (PCA). Graph based clustering was performed individually on mock and RV1A infected sample. The KNN graph was built by FindNeighbors function with first 20 principal components and k.param = 10. Louvain clustering was done using FindCluster function with resolution = 0.8 for both samples. Clusters were separately annotated in the mock and RV1A infected sample using the following markers of the major cell groups in the airway epithelium: Basal Cycling (MKI67, HIST1H4C), Basal (KRT14, KRT15, KRT5), Hillock (SPINK5, KRT13), Secretory (SCGB1A1, BPIFA1, LYPD2), Ciliated (CAPS, PIFO, MORN2), Ionocyte (FOXI1) and PNEC(pulmonary neuroendocrine cells, markers include AZGP1, AVIL)(Plasschaert et al., 2018). Each cluster found by clustering was assigned to one of the above seven major groups. A group of developing ciliated cells with marker CCNO was found in the mock sample and was merged into the ciliated cell cluster. After cluster annotation, mock and infected samples were merged together to produce tSNE maps and make comparisons. Both samples used same normalization method and the gene expression level was re-scaled. Dimensionality reduction was performed by latent semantic analysis using Seurat function RunLSI with the first 50 singular values. The tSNE maps were then produced with the first 50 dimensions and perplexity 30. The color coding of tSNE plots used cell type, sample source, viral read per cell and expression levels of genes of interest.

## KEY RESOURCES TABLE

| REAGENT or RESOURCE | SOURCE | IDENTIFIER |
| --- | --- | --- |
| **Bacterial and Virus Strains** |  |  |
| HRV-01A | ATCC | ATCC VR-481 |
| SARS-CoV-2 | BEI resources | USA-WA1/2020 |
| **Biological Samples** |  |  |
| Residual nasopharyngeal samples from clinical testing | Yale-New Haven Hospital |  |
| **Chemicals, Peptides, and Recombinant Proteins** |  |  |
| BX795 | Millipore Sigma | SML0694 |
| **Critical Commercial Assays** |  |  |
| iTaq Universal SYBR Green Supermix | BioRad | 172-5125 |
| iScript cDNA synthesis kit | BioRad | 170-8891BUN |
| Primer-probe assay for SARS-CoV-2 (N1 gene) | IDT resources | 10006600 |
| Luna Universal Probe One-Step RT-qPCR Kit | New England Biolabs | E3006L |
| QIAamp Viral RNA isolation kit | QIAGEN | 52904 |
| QIAGEN RNeasy RNA isolation kit | QIAGEN | 74106 |
| Human CXCL10/IP-10 DuoSet ELISA kit | R&D systems | DY266 |
| **Deposited Data** |  |  |
| Nasopharyngeal transcriptome data – deposited in GEO |  |  |
| Single cell RNA-Seq data – deposited in GEO |  |  |
| **Experimental Models: Cell Lines** |  |  |
| NHBE-Bronchial Epi Cells for ALI | Lonza Walkersville Inc. | CC-2540S |
| H1 HeLa cells | ATCC | ATCC CRL-1958 |
| Vero E6 cells | ATCC | ATCC CRL-1586 |
| **Oligonucleotides** |  |  |
| Primer sequences provided in Methods details |  |  |
| **Software and Algorithms** |  |  |
| R | www.r-project.org | SCR_001905 |
| Seurat | satijalab.org/seurat/ | RRID:SCR_016341 |
| Prism v. 9.0.0 for MacOS | GraphPad | RRID:SCR_005375 |
| Excel v. 16.43 | Microsoft | RRID:SCR_016137 |
| Ingenuity Pathway Analysis | QIAGEN | RRID: SCR_008653 |
| Cytoscape | Open source | RRID: SCR_003032 |

