## Supplementary material for "Magnitude and timing of the antiviral response determine SARS-CoV-2 replication early in infection": Cheemarla et al_2021_Supplemental Figures and Table

**Supplemental Figures**
**Fig S1 (related to Fig 1) Characteristics of 30 SARS-CoV-2+ patient samples used for transcriptome analysis.**

**Fig S2 (related to Fig 2). Relationship between age, sex, and NP CXCL10 in 140 SARS-CoV-2+ patients**

**Fig S3 (related to Fig 5). Expression level of full-length ACE2 and the truncated variant dACE2 in mock-treated airway epithelial organoids following rhinovirus infection (HRV-01A).**

**Fig S4. (related to Fig 5) Time course, cell type specificity, and entry receptor expression for HRV-01A in airway epithelial organoid cultures.**

**Fig S5 (related to Fig 7). Estimated doubling time for SARS-CoV-2 +/- BX795 in low MOI infection**

**Supplemental Table**

**Table S1 (related to Fig 3) Clinical description of longitudinal samples**

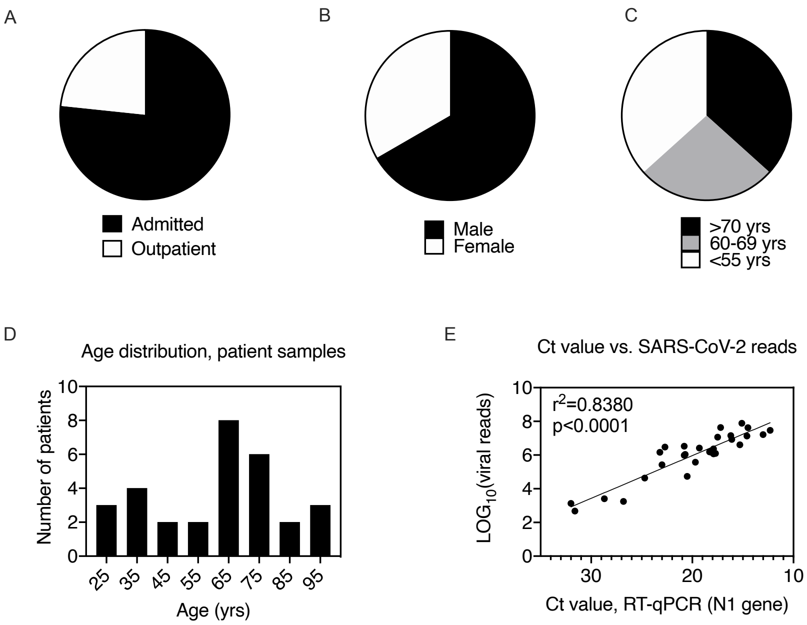

**Fig S1 (related to Fig 1). Characteristics of 30 SARS-CoV-2+ patient samples used for transcriptome analysis.**

(A-C) Patient admission status, gender, and age by category.

(D) Patient age distribution by decade. Bin center is shown on x-axis.

(E) Correlation between SARS-CoV-2 viral load measured by RT-qPCR for the viral N1 gene and number of reads mapping to SARS-CoV-2 genome by RNASeq.

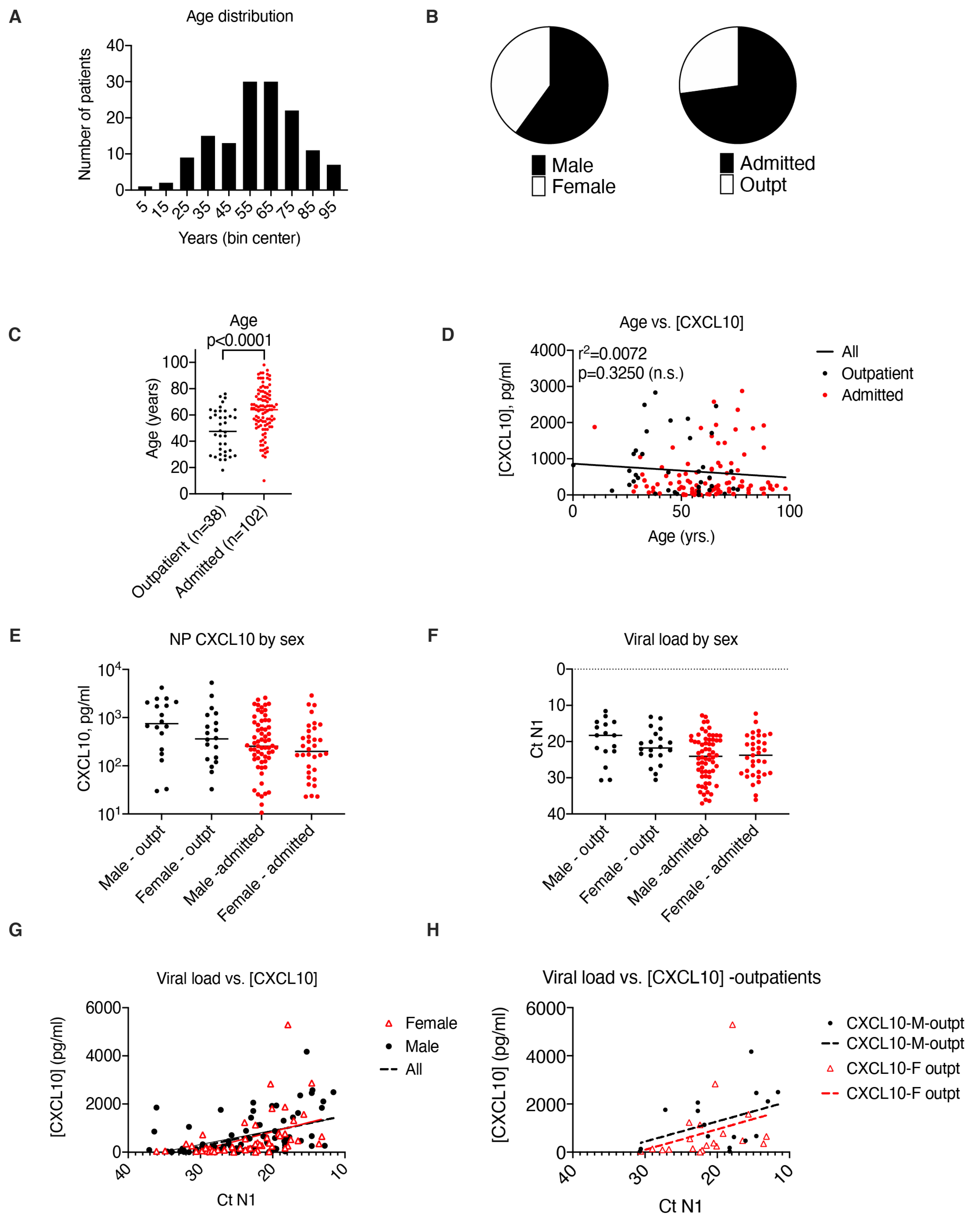

**Fig S2 (related to Fig 2). Relationship between age, sex, and NP CXCL10 in 140 SARS-CoV-2+ patients**

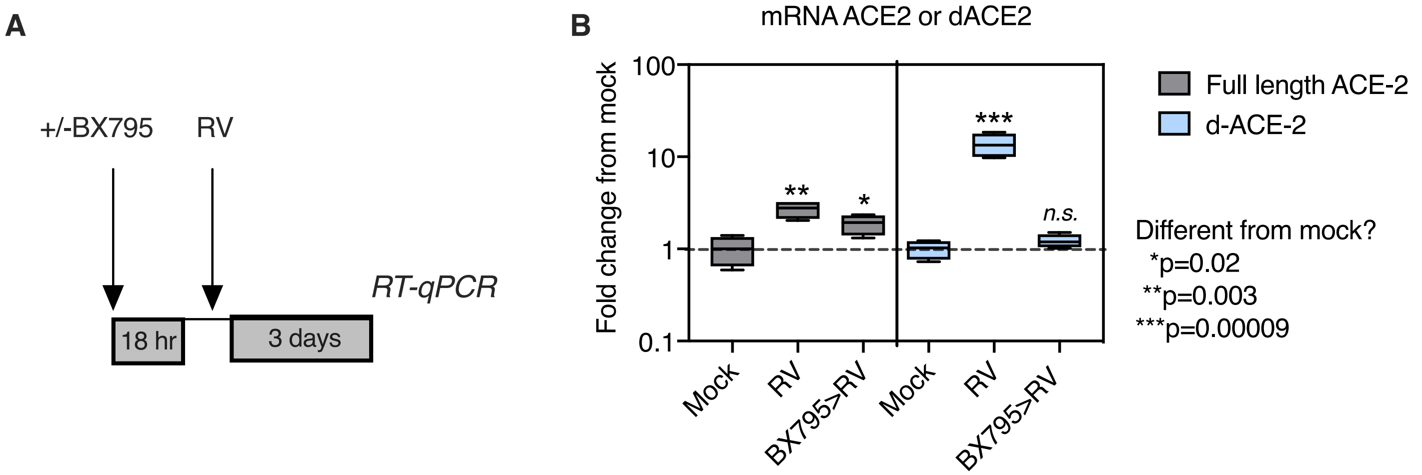

**Fig S3 (related to Fig 5). Expression level of full-length ACE2 and the truncated variant dACE2 in mock-treated airway epithelial organoids following rhinovirus infection (HRV-01A)**.

(A) Organoids were mock infected or infected with rhinovirus. Some rhinovirus-infected cultures were pre-treated for 18 hr with 6 micromolar BX795 prior to infection with rhinovirus.
(B) RT-qPCR was performed to quantify transcripts for full length ACE-2 or dACE2. Plot shows mean and range (min to max) of 4 biological replicates per condition. Asterisks indicate significant differences from mock for each condition by the student unpaired t-test. *n.s.*= not significant

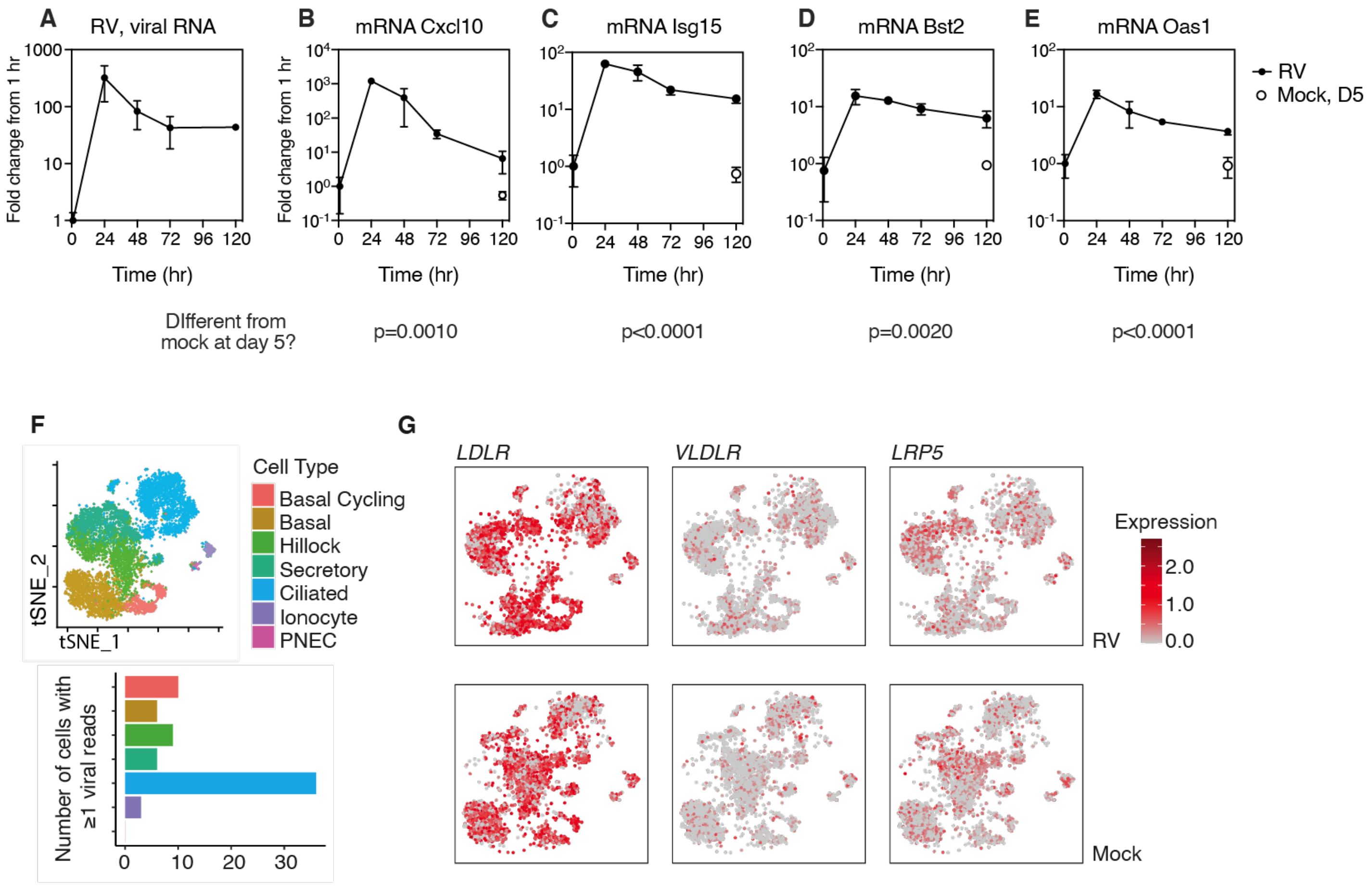

**Figure S4 (related to Fig 5). RV1A replication in differentiated primary human bronchial epithelial cultures and entry receptor expression.**

(A-E) Viral RNA level and ISG mRNA levels are graphed as fold change from ISG level at t=1 hr (post-inoculation time point). Open symbols represent ISG levels in mock-infected cultures at day 5. P-value for difference between mock and RV-infected ISG levels at day 5 are shown for each ISG (by student;s unpaired t-test.). Symbols represent mean and S.D. of 3-5biological replicates per condition.

(A) Cell type specific rhinovirus infection in human bronchial epithelial organoid cultures, 5 days post infection with rhinovirus (HRV-01A) or mock-infection. Top panel shows cell types present; bottom panel shows cell type specific distribution of 70 cells containing at least one viral read.

(B) Expression level of entry receptors for HRV-01A (LDLR, VLDLR, LRP5), in rhinovirus infected (top) or in mock-treated cultures (bottom panels.)

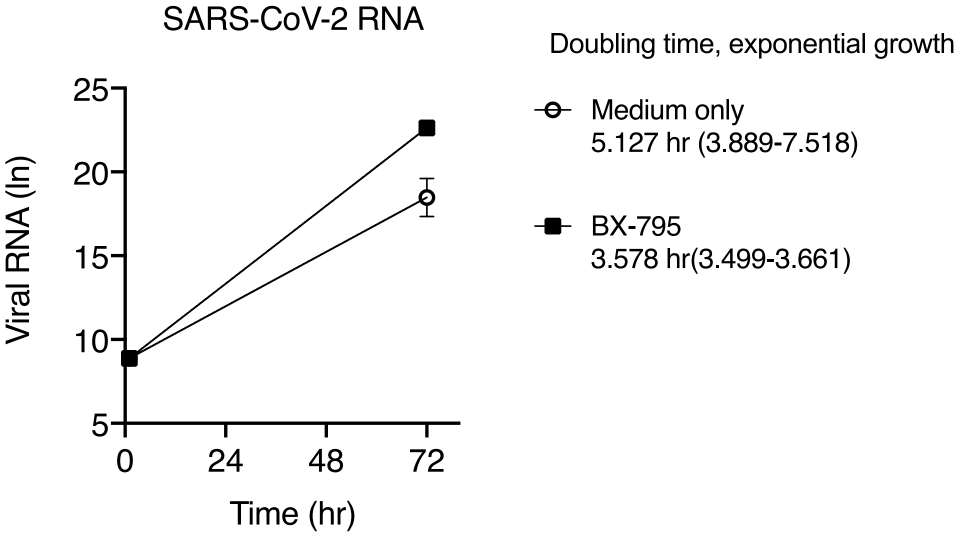

**Fig S5 (related to Fig 7). Estimated doubling time for SARS-CoV-2 +/- BX795 in low MOI infection**

### Table S1 (related to Fig 3). Clinical information for longitudinal patients

| Patient | Age | Sex | Presentation | Clinical course | NP CXCL10, pg/ml | Ct value, N1 |
|---|---|---|---|---|---|---|
| **Viral load low to high** | | | | | **First sample in series** | |
| **Patient** | **Age** | **Sex** | **Presentation** | **Clinical course** | **NP CXCL10, pg/ml** | **Ct value, N1** |
| L2 | 60s | M | screen+, asymptomatic | no SARS-CoV-2 symptoms | not detected | 34.6 |
| L12 | 50s | M | screen+, asymptomatic | no SARS-CoV-2 symptoms | 31 | 31.9 |
| L39 | 70s | F | fever | moderate course, no ICU | 66 | 30.7 |
| L42 | 80s | F | fever and cough | no oxygen requirement, no ICU | 103 | 32.8 |
| L5 | 80s | M | fever, cough, dyspnea | moderate course, no ICU | 137 | 29.4 |
| L44 | 90s | F | fever and cough | no oxygen requirement, no ICU | 152 | 28.7 |
| **Viral load high to low** | | | | | **First sample in series** | |
| **Patient** | **Age** | **Sex** | **Presentation** | **Clinical course** | **NP CXCL10, pg/ml** | **Ct value, N1** |
| L8 | 80s | F | confusion, hypoxia | moderate course, no ICU | 1745 | 18.6 |
| L45 | 90s | F | fever, dyspnea | moderate course, no ICU | 452 | 17.4 |
| L18 | 70s | F | dyspnea | moderate course, no ICU | not available | 15.5 |
| L1 | 60s | M | cough, hypoxia | moderate course, no ICU | 675 | 14.8 |
| L30 | 70s | F | fever, dyspnea | moderate course, no ICU | 3109 | 14.6 |
| L23 | 90s | F | screen+, asymptomatic | no oxygen requirement, no ICU | 4602 | 12.6 |
| L41 | 70s | M | cough | no oxygen requirement, no ICU | 3270 | 10.8 |
